# Investigating the healthcare-seeking behaviors of mobile phone users in rural Uganda

**DOI:** 10.1101/2023.10.30.23297770

**Authors:** Nelly Mwandacha, Hallie Dau, Maryam AboMoslim, Priscilla Naguti, Mia Sheehan, Amy Booth, Laurie Smith, Jackson Orem, Gina Ogilvie, Carolyn Nakisige

## Abstract

Cervical cancer is the leading cause of cancer in low- and middle-income countries, despite being a preventable disease. Uganda, which lacks an effective screening program, has one of the highest cervical cancer incidence rates in the world. Mobile health (mHealth) technology has the potential to improve healthcare-seeking behaviors and access to cervical cancer screening. This study aims to describe the connection between mobile phone access and healthcare-seeking behaviors in rural Uganda.

This cross-sectional study recruited participants from January 23 to August 24, 2023. Women were eligible if they had no prior screening or treatment for cervical cancer in the past 5 years, were aged 30 to 49 years old, and were residents of the South Busoga Forest reserve. Each participant completed a 43-item survey which included questions on demographics, previous health service usage, and opinions on cervical cancer prevention. All data was analyzed using descriptive statistics and chi-square tests.

Of the 1434 participants included in the analysis, 91.4% reported having access to a mobile phone. Most respondents were aged 30-40 years, were married or in a relationship, had ≤ primary education, and were farmers. Participants with access to a mobile phone were significantly more likely to report attending a healthcare outreach visit (access = 87.3%, no access = 72.6%, p<0.001) or visiting a health centre (access = 96.9%, no access = 93.5%, p<0.001). Participants in both groups had largely positive attitudes around and good knowledge of cervical cancer screening.

While attendance to healthcare outreach visits or health centres was high amongst participants, those with mobile phone access were more likely to seek healthcare services. Further inquiry into this association between mobile phone access and healthcare-seeking behaviour is needed to optimize the improvements to cervical cancer screening when implementing interventions such as mHealth technology.

**AUTHOR SUMMARY:** Cervical cancer is the leading cause of cancer in low- and middle-income countries, despite being a preventable disease. This can be partially attributed to the lack of widespread screening programs. In Uganda, the development of a comprehensive screening program has been slow despite having one of the highest rates of cervical cancer. However, mobile health might have the capacity to help improve cervical cancer screening rates in resource-limited settings such as Uganda. Our study explored the existing relationships between access to a mobile phone and healthcare-seeking behaviour in rural Uganda. We found that access to a mobile phone was associated with higher use of healthcare services and a more positive attitude towards and knowledge of cervical cancer prevention. It is important to study these existing relationships to find the best use of mobile health and to allow for the assessment of a digital health intervention once implemented. Future studies can build on our findings by investigating the impact of digital health interventions on the use of cervical cancer screening services in rural settings, which will contribute to the elimination of this devastating disease in Uganda, and in other resource-limited settings.

## INTRODUCTION

Cervical cancer is the second most common cancer affecting women in low- and middle-income countries (LMICs) [1]. In 2020 alone, over 600,000 cases of cervical cancer were reported worldwide [2]. While largely considered a preventable cancer, immense disparities in the burden of the disease remain, with the highest proportions of morbidity and mortality concentrated in LMICs [2]. The high rates of cervical cancer in LMICs can be attributed to inadequate screening [1,2]. Uganda has one of the highest cervical cancer incidence rates in the world, with a rate three-times higher than the global average [3,4]. The development of a comprehensive screening program in Uganda, which would alleviate the cervical cancer burden, has been hampered by economic and accessibility constraints including a lack of infrastructure and trained personnel [3–5]. As a result, cervical cancer in Uganda is often diagnosed at an advanced stage [3,4]. As such, there is a need to better understand how to develop a more widespread, effective, and accessible cervical cancer screening program in Uganda and similar LMICs.

Healthcare-seeking behaviour, defined as the actions an individual takes when they self-identify as needing a health service or being in poor health, plays a critical role in the final decision to interact with health services [6–8]. For cervical cancer screening, lower healthcare-seeking behaviour overall often translates into lower intention to screen, and may result in lower cervical cancer screening uptake [9]. Importantly, healthcare-seeking behaviour has been found to be influenced by several factors such as community norms, provider availability, service costs and education [6,10–13]. Previous inquiries into the factors influencing healthcare-seeking behaviour in Uganda have implicated cost of services, long travel times, income, and education as impactful factors [14,15]. However, few studies in Uganda have probed the relationship between mobile phone access and healthcare-seeking behaviour.

Mobile health (mHealth) technology, defined as the facilitation of healthcare or public health through mobile devices, has emerged as a promising tool to facilitate healthcare service usage and improve health outcomes in hard-to-reach, underserved populations [16]. Mobile phone usage has grown immensely in Uganda in recent decades with 71% of Ugandans reporting mobile phone ownership in a 2018 report [17]. Due to this widespread reach, mobile phones have been identified as a promising intervention for the delivery of health information and services in Uganda and other countries in Eastern Africa [18–20]. For instance, Huchko et al. found in their 2019 cluster-randomized trial that women in rural Kenya with improved healthcare-seeking behaviour, such as increased usage of family planning and HIV testing services, preferred receiving cervical cancer screening results via their mobile phone over receiving results through a home visit [21]. However, there is a gap in understanding how this technology can be used in Uganda to improve healthcare seeking-behaviour, including cervical cancer screening uptake, particularly in rural regions which have limited access to screening services [22]. As such, to optimize healthcare utilization in rural Uganda through the introduction of mHealth technology and facilitate the evaluation of these interventions, it is imperative to first investigate existing associations between access to technology and healthcare-seeking behaviour. Consequently, the objective of this analysis is to describe the connection between mobile phone access and healthcare-seeking behaviors in rural Uganda.

## METHODS

### Design, Setting and Study Population

This cross-sectional analysis utilizes survey data collected as part of a pragmatic trial in Malongo, an extremely rural sub-county of the Mayuge District in Eastern Uganda. The survey was administered to eligible women at baseline, after their enrollment in the trial. Women were eligible for this analysis if they had no prior screening or treatment for cervical cancer in past 5 years, were aged 30 to 49 years old, and were residents of one of the eleven selected villages in the South Busoga Forest reserve. Those who were pregnant, had a hysterectomy, and/or were unable to provide informed consent were excluded from the study. Recruitment into the study was done by trained village health teams (VHTs) through door-to-door home visits. VHTs collected survey data digitally on tablets using REDCap [23,24], a secure electronic data collection system. To ensure inclusion of women with low literacy levels, surveys were administrated orally by VHTs in Lusoga or English. Data collection started in January 23, 2023 and was completed in August 24, 2023.

### Survey

The data in this analysis comes from a baseline survey that consisted of 43 main questions and 16 follow-up questions. It aimed to determine medical history and assess knowledge and attitudes surrounding cervical cancer and HPV prior to the intervention. The survey included questions on participant demographics, previous health services usage, and opinions on cervical cancer prevention and screening.

To determine mobile phone access, women were asked if they had access to a mobile phone, whether that be their own or through family, friend, or neighbour. Women had the option to select yes (‘access’), no, or don’t know. No and don’t know options were combined and designated as not having mobile phone access (‘no access’).

Healthcare-seeking behaviour was characterized as the outcome of this analysis and was conceptualized using several questions. Women were asked if they had ever attended a healthcare outreach visit, locales where otherwise absent services are brought into a community on a temporary or mobile basis, or a health centre, permanent community clinics that offer primary and preventive care. The available answer options were yes, no, or don’t know with follow-up questions [25,26]. The survey assessed attitudes surrounding cervical cancer prevention and curability. Women were asked about the importance of early detection, the curability of cervical cancer following early detection, vaccinations against cervical cancer and, ultimately, if cervical cancer can be prevented. Response options for these questions were yes, no, don’t know, or refused. Finally, women were asked about their choice to receive an HPV vaccine if one was available for their age group; participants answered on a five-point Likert scale, with response options varying from strongly agree to strongly disagree. Responses were recategorized to be dichotomous in which strongly agree and somewhat agree were labelled as yes and all other responses were recategorized as no.

In addition to the questions related to the exposure and outcome described above, the survey asked several demographic questions that were utilized to determine the characteristics of the women participating in the study. These included age, number of pregnancies, marital status, education level, current partner’s education level (if applicable), and HIV status.

### Statistical Analysis

Analysis of the relevant survey questions was conducted using R version 4.3.0 [27] and R Studio [28]. Bivariate statistical analyses were conducted using chi-squared tests to compare the outcomes of those with mobile phone access and those without mobile phone access. P-values < 0.05 were considered statistically significant [29]. All missing data was included in the results.

### Ethics

This study posed minimal risk to the participants. Ethics approval was obtained from the University of British Columbia Children and Women’s Research Ethics Board (H22-01634), Uganda Cancer Institute Research Ethics Committee (UCI REC) (UCI-2022-56), and the Uganda National Council for Science and Technology (UNCST). Participants were compensated 20,000 UGX upon completion of the screening study procedures, which includes the baseline survey.

## RESULTS

In total, 1434 participants were included in the analysis. A majority of participants, 1310 (91.4%), reported having access to a mobile phone, while 124 (8.6%) reported not having access to a mobile phone. Most respondents were between the ages of 30 and 40 years old (n = 929), reported being married or in a relationship (access, n = 1133, 86.5%; no access, n = 106, 85.5%; p= 0.948), had completed primary education or less (access, n = 1157, 88.3%; no access, n = 114, 91.9%; p=0.434), and reported farming as their occupation (access, n = 1114, 85.0%; no access, n = 101, 81.5%; p=0.438). A significantly larger proportion of women with HIV (n=125) were in the “no access” group (access, n = 102, 7.8%; no access, n = 23, 18.5%; p<0.001). Additional demographic characteristics are detailed in **Table 1**.

**Table 1.** Demographics of participants with and without mobile phone access.

|  |  | **Total**<br>n = 1434<br>n (%) | **Yes**<br>n = 1310<br>n (%) | **No**<br>n = 124<br>n (%) | **p-value** |
| --- | --- | --- | --- | --- | --- |
| **Mobile Phone Access** | Yes | 1310 (91.4) | 1310 (100.0) | 0 (0.0) | <0.001 |
|  | No | 124 (8.6) | 0 (0.0) | 124 (100.0) |  |
| **Language** | English | 37 (2.6) | 30 (2.3) | 7 (5.6) | 0.05 |
|  | Lusoga | 1397 (97.4) | 1280 (97.7) | 117 (94.4) |  |
| **Age** | 30-34 | 611 (42.6) | 556 (42.4) | 55 (44.4) | 0.973 |
|  | 35-39 | 318 (22.2) | 291 (22.2) | 27 (21.8) |  |
|  | 40-44 | 249 (17.4) | 229 (17.5) | 20 (16.1) |  |
|  | 45-49 | 256 (17.9) | 234 (17.9) | 22 (17.7) |  |
| **Marital Status** | Married/In a relationship | 1239 (86.4) | 1133 (86.5) | 106 (85.5) | 0.948 |
|  | Single | 116 (8.1) | 106 (8.1) | 10 (8.1) |  |
|  | Separated/<br>Divorced/<br>Widowed | 78 (5.4) | 70 (5.3) | 8 (6.5) |  |
|  | *Missing* | 1 (0.1) | 1 (0.1) | 0 (0.0) |  |
| **Education** | ≤ Primary school | 1271 (88.6) | 1157 (88.3) | 114 (91.9) | 0.434 |
|  | > Primary school | 153 (10.7) | 144 (11.0) | 9 (7.3) |  |
|  | *Missing* | 10 (0.7) | 9 (0.7) | 1 (0.8) |  |
| **Partner Education** | ≤ Primary school | 1004 (70.0) | 914 (69.8) | 90 (72.6) | 0.602 |
|  | > Primary school | 218 (15.2) | 203 (15.5) | 15 (12.1) |  |
|  | *Missing* | 212 (14.8) | 193 (14.7) | 19 (15.3) |  |
| **Occupation** | Farmer | 1215 (84.7) | 1114 (85.0) | 101 (81.5) | 0.438 |
|  | Other | 177 (12.3) | 156 (11.9) | 21 (16.9) |  |
|  | *Missing* | 42 (2.9) | 40 (3.1) | 2 (1.6) |  |
| **Number of Pregnancies** | 0-5 | 38 (2.6) | 35 (2.7) | 3 (2.4) | 0.573 |
|  | 6-10 | 219 (15.3) | 204 (15.6) | 15 (12.1) |  |
|  | >10 | 1177 (82.1) | 1071 (81.8) | 106 (85.5) |  |
| **Self-Reported Positive HIV Test** | Yes | 125 (8.7) | 102 (7.8) | 23 (18.5) | <0.001 |
|  | No | 1302 (90.8) | 1202 (91.8) | 100 (80.6) |  |
|  | *Missing* | 7 (0.5) | 6 (0.5) | 1 (0.8) |  |

Among the women surveyed, 1143 (87.3%) of individuals in the access group and 90 (72.6%) of those in the no access group reported ever attending a healthcare outreach visit (p<0.001). Additionally, 96.9% (n = 1269) of the access group and 93.5% (n = 116) of the no access group reported that they had ever visited a health centre (p<0.001).

Women in both groups were knowledgeable about cervical cancer prevention. 1051 (80.2%) in the access group and 104 (83.9%) in the no access group identified that cervical cancer can be prevented (p=0.656). Moreover, 1188 (90.7%) in the access group and 111 (89.5%) in the no access group noted that that early detection of cervical cancer is important (p=0.427). Similarly, 1075 (82.1%) in the access group and 107 (86.3%) in the no access group correctly recognized that cervical cancer is curable if detected early (p=0.035). Finally, most women in both groups identified that one can be vaccinated against cervical cancer (access, n = 1042, 79.5%; no access, n = 101, 81.5%; p=0.504). Notably, there was a resoundingly positive response to inquiries about the acceptance of a preventative HPV vaccine in which 1309 (99.9%) women in the access group and 123 (99.2%) of women in the no access group agreed they would take a vaccine for their age group (p=0.410).

## DISCUSSION

This analysis evaluated the associations between mobile phone access and healthcare-seeking behaviour of women in the rural, South Busoga Forest reserve in the Malongo sub-county of Uganda. We found that, while the majority of both the access group and the no access group reported having attended a healthcare outreach visit or visited a health centre, a greater percentage of women in the access group had sought out these services. This finding is consistent with studies that have looked at mobile phone ownership and its impacts on reproductive and sexual health-related visits and behaviors [30]. For example, in a 2020 inquiry into the health outcomes of 15 LMICs including Uganda, LeFevre et al. found an association between mobile phone ownership and improved attendance to antenatal care clinic visits, increased uptake of vaccines during pregnancy, and improved post-natal care for both women and newborns [30]. Access to a mobile phone appears to be associated with increased usage of healthcare services and, as such, improved healthcare-seeking behaviour.

Of women who had exhibited healthcare-seeking behavior in the past, women in both access groups were more likely to have visited a health centre than have attended a healthcare outreach visit. Higher attendance at health centre could be due to the fact that residents are more likely to seek out services, typically at a health centre, when needed; women are often required to travel outside of their communities, as opposed to waiting for an outreach initiative to come to the community [22]. Due to the urban-rural health infrastructure disparities in Uganda, women in remote areas such as Malongo are often required to engage in active healthcare-seeking behaviour to receive the necessary health services rather than passively attending outreach visits when available. Additionally, health centres are available continuously throughout the year while health outreach visits are established for temporary periods of time, potentially resulting in a greater percentage of respondents visiting health centres [25].

Women in both the access and no access groups predominantly had positive attitudes towards cervical cancer prevention and screening. This finding is both consistent with studies that have assessed attitudes surrounding cervical cancer and contradictory to studies that have assessed knowledge of the disease conducted in both urban and rural settings in Uganda [31–35]. In their 2017 cross-sectional study in the Mayuge and Bugiri districts of eastern Uganda, Mukama et al. found knowledge and attitudes around cervical cancer screening and vaccination to be mainly positive amongst the women surveyed [31]. Similarly, a 2017 qualitative study conducted by Turiho et al. in western Uganda found largely favourable opinions of the HPV vaccine amongst young girls, their parents and communities, highlighting similar support for cervical cancers preventative measures as was found in our analysis [32]. This recognition of the importance of cervical cancer screening and prevention amongst women in Uganda is encouraging as these positive attitudes could manifest in increased support for and participation in preventative interventions. Given the low screening rates throughout the country that are often attributed to accessibility and economic constraints, these positive attitudes, when paired with interventions designed to address these limitations, could improve the cervical cancer screening uptake in the region. Contrastingly, Ndejjo et al., in their 2017 qualitative study in the Mayuge and Bugiri disticts of eastern Uganda, found most participants had limited knowledge of cervical cancer prevention and had many misconceptions surrounding causes of cervical cancer [34]. Similarly, a 2017 northern Ugandan cross-sectional study conducted by Waiswa et al. found lower knowledge of cervical cancer prevention and screening amongst participants [35]. These differences in results demonstrated in previous literature could be attributed to drastic differences in study design. Nonetheless, our study findings are promising due to the importance of prevention and early screening to positive cervical cancer outcomes [4].

Out study was strengthened by the employment of VHTs as they provide community engagement, sustainability, and cultural sensitivity as participants can respond in their preferred language to a fellow community member [36]. Furthermore, the study team consisted of a diversity of individuals with extensive knowledge of cervical cancer and experience conducting this type of research. However, as individuals self-reported responses, recall bias could have influenced the results and is considered a limitation [37]. In addition, respondents delivered responses to survey questions to VHTs verbally which could result in social desirability bias [37]. Moreover, this study was conducted in exclusively in the Malongo sub-county and may not represent experiences throughout the country or other rural LMIC settings. Finally, knowledge of the extent and amount of mobile phone usage and access by the women surveyed was limited in this study. Digital illiteracy, limited availability of chargers and electricity, expensive data and airtime costs, and unequal phone sharing have been documented throughout Uganda, especially in remote regions of the country [38–40]. Consequently, it was assumed that mobile phone access equated to utilization in this inquiry.

While knowledge and attitudes surrounding cervical cancer prevention are largely supportive, a gap remains between these positive opinions and healthcare service uptake. The findings of our analysis suggest a need for interventions that improve cervical cancer accessibility and availability issues in rural Uganda. As access to healthcare services in rural regions such as Malongo remains limited, future research could investigate the potential impacts of ‘take-home’ screening methods such as HPV-based self-collection for cervical cancer screening. HPV-based self-collection minimizes the staff and infrastructure required for screening and mitigate discomfort and privacy concerns surrounding traditional screening methods that require pelvic exams [41,42]. However, concerns surrounding the mechanisms of screening information and results delivery with HPV-based self-collected screening prevail; mHealth interventions to facilitate screening information and results delivery via mobile phones which would address this limitation [42–44].

Our analysis demonstrates that, while most women in the rural Malongo have attended a healthcare outreach visit or a health centre, women with mobile phone access were more likely to utilize these services. Despite these differences, both women with and without mobile phone access had positive attitudes surrounding and knowledge of cervical cancer prevention. By continuing these inquiries into facilitators to increase uptake of screening and prevention measures, we can continue making progress towards the elimination of cervical cancer, both in Uganda and other LMICs.

## Data Availability

Data is not available for access as it contains human data that contains potentially identifying information about participants. All data inquires can be made to the UBC Research Ethics Board (contact via).

